# Aggregated eosinophils characterize airway mucus properties

**DOI:** 10.1101/2022.11.15.22282331

**Authors:** Yui Miyabe, Mineyo Fukuchi, Hiroki Tomizawa, Yuka Nakamura, Mitsutoshi Jikei, Yoshinori Matsuwaki, Misaki Arima, Yasunori Konno, Yuki Moritoki, Masahide Takeda, Naoya Tanabe, Hiroshi Sima, Yusuke Shiraishi, Toyohiro Hirai, Nobuo Ohta, Junko Takahata, Atsushi Matsubara, Takechiyo Yamada, Koichiro Asano, Isao Miyairi, Rossana C. N. Melo, Peter F. Weller, Shigeharu Ueki

## Abstract

Uncontrolled airway mucus is associated with diverse diseases. We hypothesized that the physical characteristics of infiltrating granulocytes themselves affect the clinical properties of mucus. Surgically obtained nasal mucus from patients with eosinophilic chronic rhinosinusitis (ECRS) and neutrophil-dominant non-eosinophilic chronic rhinosinusitis (non-ECRS) was assessed in terms of computed tomography (CT) density, viscosity, water content, wettability, and granulocyte-specific proteins. In an observational study, we found that nasal mucus from patients with ECRS had significantly higher CT density, viscosity, dry weight, and hydrophobicity than mucus from patients with non-ECRS. The levels of eosinophil-specific proteins in nasal mucus correlated with its physical properties. When isolated human eosinophils and neutrophils were stimulated to induce extracellular traps followed by aggregate formation, we found that cell aggregates showed physical and pathological findings that closely resembled mucus. Co-treatment with heparin (which slenderizes the structure of eosinophil extracellular traps) and DNase efficiently induced a reduction in the viscosity and hydrophobicity of both eosinophil aggregates and eosinophilic mucus. The present study highlights the pathogenesis of mucus stasis in infiltrated granulocyte aggregates from a new perspective. The combination of DNase and heparin might be a novel therapeutic modality against pathologic viscous eosinophilic mucus.

**One Sentence Summary:** Intraluminal accumulation and activation of eosinophils contribute to the clinical properties of airway mucus and may serve as a therapeutic target.

## INTRODUCTION

Airway mucus has a role as a physical barrier to prevent the entry of external pathogens and contributes to the expulsion of foreign substances through mucociliary transport and maintenance of local humidity *(1)*. However, abnormal production, hypercondensation, and decreased clearance of airway mucus can lead to airway obstruction and prolonged inflammation, thereby leading to a poor prognosis *(2)*.

In bacterial infection, airway mucus contains abundant neutrophils that migrate into the airways to eliminate pathogens through phagocytosis. These neutrophils also possess anti-microbial capacity in the form of the release of sticky chromatin structures, i.e., neutrophil extracellular traps (NETs). NETs serve as scaffolding for antimicrobial mediators such as histones and granule proteins, and contribute to the entrapment of bacteria and other extracellular pathogens *(3)*. Abundant eosinophil extracellular traps (EETs) were also observed in patients with diverse eosinophilic diseases *(4, 5)*. These extracellular traps (ETs) are major components of inflammatory cell-rich mucus, and contribute to its viscosity.

Interestingly, a clinical characteristic of eosinophil-dominant mucus, which is often associated with type 2 inflammation, is that it shows high viscosity. For instance, eosinophilic chronic rhinosinusitis (ECRS) is resistant to conservative treatments using antimicrobial therapies or nebulizers, and frequently requires surgical intervention for mucus removal, which is in contrast to infectious rhinosinusitis *(6, 7)*. Luminal impaction due to viscous mucus is also observed in other eosinophilic upper and lower airway diseases such as eosinophilic otitis media *(5)*, allergic fungal sinusitis *(8)*, allergic bronchopulmonary aspergillosis *(9-11)*, plastic bronchitis *(12, 13)*, and eosinophilic asthma *(14-16)*. Eosinophilic airway mucus has been clinically recognized as allergic mucin, eosinophilic mucin, and mucus plug, and its consistency has been described as being like chewing gum, cottage cheese, axle grease, and peanut butter, which can be a clue to the diagnosis *(17-19)*. The clinical similarity of eosinophil-rich mucus in different airway diseases has been previously discussed *(20)*, although the precise mechanisms leading to its accumulation are less well understood.

It is of paramount importance to elucidate the mechanisms and appropriate control of mucus in muco-obstructive airway diseases. The objective of this study was to clarify the contribution of granulocytes to the characteristics of pathological mucus. Using multiple modalities including biophysical approaches, our results highlight the differences between neutrophil- and eosinophil-rich mucus, and the contributions of aggregated extracellular trap-producing granulocytes.

## RESULTS

### Eosinophilic mucus shows high CT density

Microscopically, mucus from patients with ECRS is characterized by the presence of abundant eosinophil accumulations, whereas that from non-ECRS patients is characterized by neutrophil accumulations (**Fig. 1A**). First, we questioned whether, in addition to differences in cellular composition, mucus density would also vary between ECRS and non-ECRS patients. In typical eosinophilic mucus, a unique radiological imaging presentation is high attenuation on computed tomography (CT) *(10, 21)*. Accordingly, as shown in **Fig. 1B**, the mucus in patients with ECRS often shows a high CT attenuation area, unlike that in non-ECRS patients. Surgically obtained mucus was hyperconcentrated as shown by macroscopic images in **Fig. 1C** and **Movie S1**. Mucus collected from patients with ECRS was typically highly viscoelastic and thicker than that from patients with non-ECRS. We initially aimed to quantify the CT value of mucus on routine sinus CT imaging; however, mucosal thickening and mucus were indistinguishable. To solve this problem, we conducted an observational study to collect mucus samples from patients with ECRS and non-ECRS who needed surgical treatment, and established *ex-vivo* CT imaging (**Fig. 1D, Fig. S1**). The patient demographics are indicated in **Table S1**. As expected, the mean CT value of mucus from patients with ECRS showed significantly higher x-ray attenuation than mucus from patients with non-ECRS (48.5 ± 16.9 Hounsfield units [HU] vs 34.2 ± 6.9 HU, p<0.01, **Fig. 1D**). These results indicate the presence of radiographical differences between eosinophil- and neutrophil-dominant mucus.

**Fig. 1.**
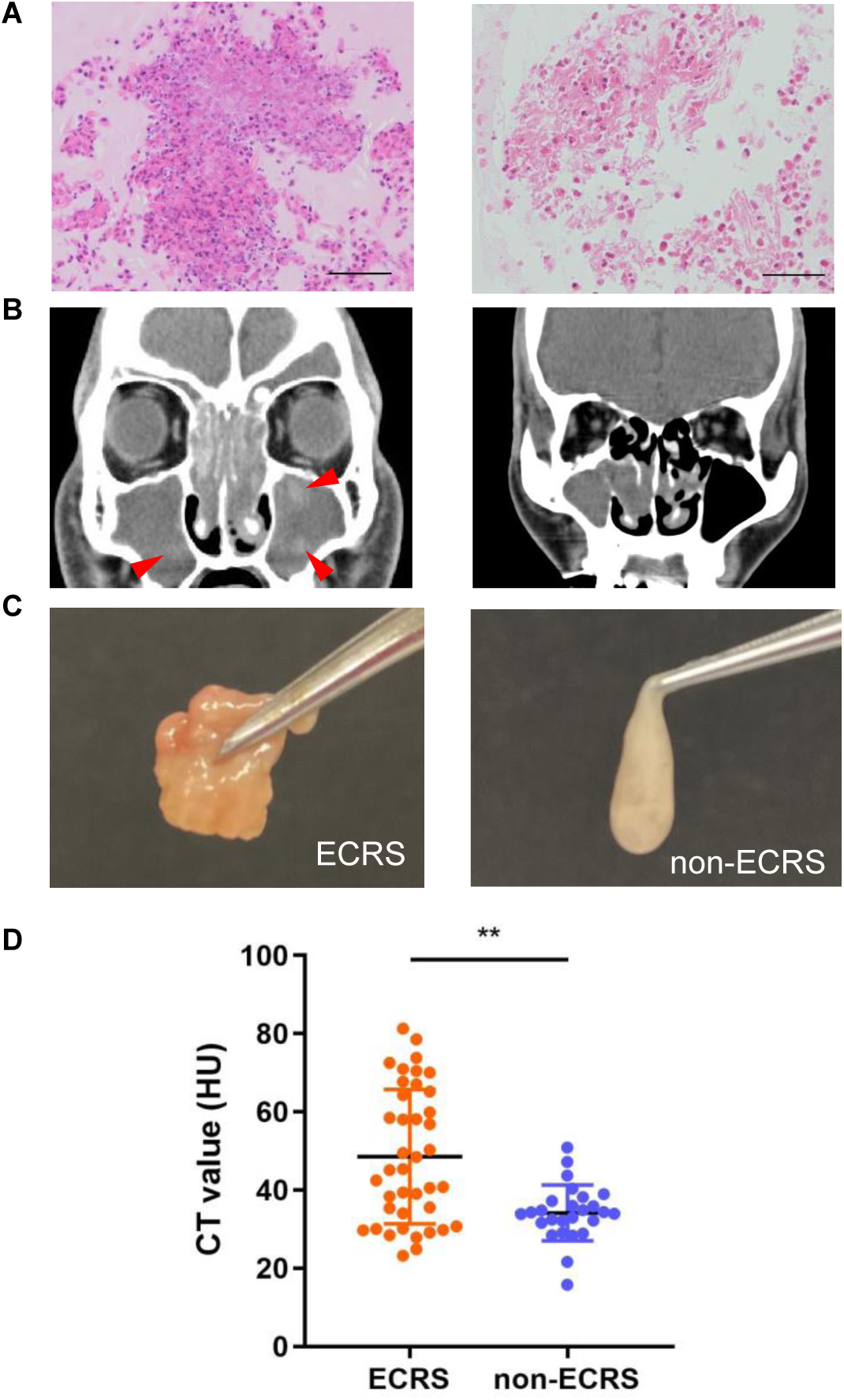
Eosinophil-rich mucus from patients with ECRS showed high CT x-ray attenuation. **(A)** The microscopic appearance of sectioned mucus in patients with ECRS and non-ECRS (400×, hematoxylin-eosin staining, scale bar=50 μm). (**B**) Coronal sinus CT images of patients with ECRS (right) and non-ECRS (left). Red arrowheads indicate high CT attenuation areas in the maxillary sinus. (**C**) Macroscopic images of surgically-obtained sinus mucus from patients with ECRS and non-ECRS. (**D)** Comparison of mucus CT values between ECRS and non-ECRS groups (ECRS: n=40; non-ECRS: n=26, mean ± SD, **p<0.01, Welch’s *t* test).

### Biophysical properties of mucus are associated with its CT density

Next, we assessed the rheological properties of mucus samples, measuring their shear viscosity using a rotational viscometer (**Fig. 2A**). With the plate in contact with the mucus sample, the shear viscosity was measured at different values of shear stress. As shown in **Fig. 2B**, the mucus viscosity decreased when the shear rate increased, indicating a non-Newtonian shear-thinning pattern, as found in previous studies on airway mucus *(22)*. As expected, higher shear viscosities were obtained in the mucus from patients with ECRS than in that obtained from patients with non-ECRS. A positive correlation between viscosity and CT values was also observed **(Fig. 2C)**. Next, we assessed the dry weight of the mucus using a thermogravimetric analyzer (TGA) **(Fig. 2D)**. The weight reduction of ECRS mucus took a longer time than non-ECRS mucus to reach a plateau when the mucus samples were concentrated by gradually heating up to 100°C **(Fig. 2E)**, indicating a higher polymer concentration. In a randomly selected sample (because of technical limitations) that we tested, the dry weight of mucus from patients with ECRS was significantly higher than that of patients with non-ECRS (40.6 ± 7.0% vs 19.4 ± 6.2%, p<0.01), and positively correlated with the CT value **(Fig. 2F)**. To further determine the wettability of the mucus, the air-dried mucus surface was analyzed with a static water contact angle **(Fig. 2G)**. The mucus from patients with ECRS showed significantly higher hydrophobicity than that from patients with non-ECRS (75.5 ± 18.3° vs 44.9 ± 18.6°, p<0.0001, **Fig. 2H)**. The mucus-water contact angles and CT values were positively correlated **(Fig. 2I)**. These results indicated that the biophysical properties of the mucus were associated with its radiographical findings.

**Fig. 2.**
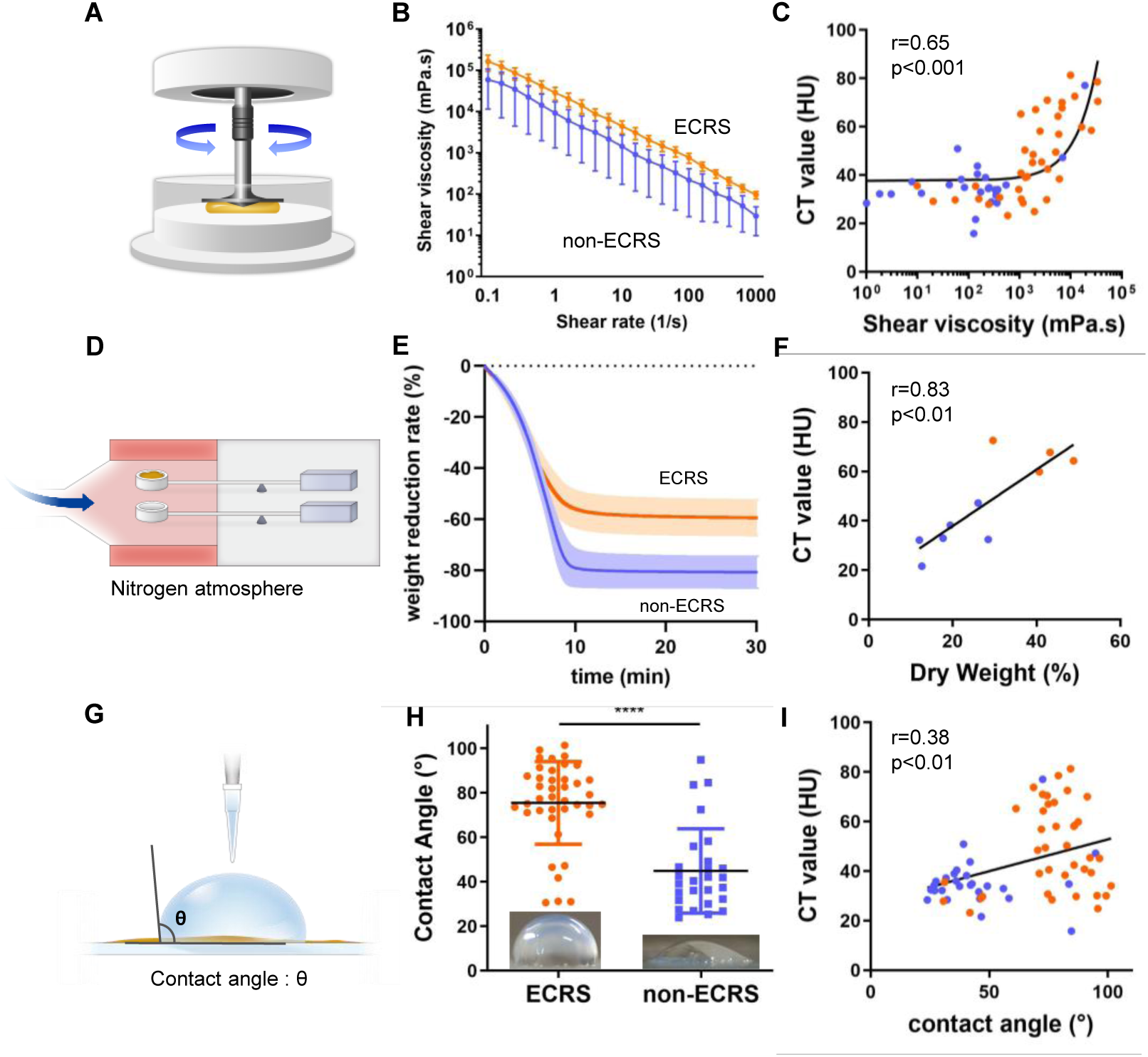
Biophysical properties of mucus and their associations with CT density. **(A)** Measurement of dynamic viscosity against the shear rate of mucus samples using a rotational viscometer. **(B)** The change in shear viscosity according to the shear rate (orange line: ECRS, n=27; blue line: non-ECRS, n=15). **(C)** Correlation between shear viscosity at a shear rate of 10 s^-1^ and the CT values of each sample (orange: ECRS, n=37; blue: non-ECRS, n=25). Coefficient of correlation, *R*^2^□ = 0.45 **(D)** Measurement of dry weight of the mucus using a thermogravimetric analyzer (TGA). **(E)** The weight reduction rate of the sample measured using TGA. Samples were gradually heated from 0 to 7.5 minutes and then kept at 100°C. Lines and colored areas indicate mean and ± SD, respectively (orange: ECRS, n=4; blue: non-ECRS, n=6). **(F)** Correlation between dry weight and CT values (orange: ECRS, n=4; blue: non-ECRS, n=6). Coefficient of correlation, *R*^2^□=□0.69 **(G)** Measurement of the contact angle (θ) between water and mucus surface. **(H)** Comparison of contact angle (orange: ECRS, n=40; blue: non-ECRS, n=27, ****p<0.0001, Mann-Whitney U test). **(I)** Scatter plot shows correlation between contact angle and CT values of the samples (orange: ECRS, n=40; blue: non-ECRS, n=27). Coefficient of correlation, *R*^2^□= □0.15.

### Eosinophilic-specific protein contents correlate with the biophysical properties of mucus

To determine whether the biophysical properties of the mucus were associated with the eosinophils in it, we measured the concentrations of the eosinophil-specific proteins galectin-10 and eosinophil-derived neurotoxin (EDN). The concentrations of galectin-10 (**Figure 3A**) and EDN (**Figure 3B**) were significantly higher in mucus from patients with ECRS than in that from patients with non-ECRS (galectin-10, 0.26 ± 0.06 vs 0.008 ± 0.005, p<0.01; EDN, 2.09 ± 0.16 vs 1.25 ± 0.12 µg/ml, p<0.001). In contrast, the concentration of neutrophil granule protein myeloperoxidase (MPO) was significantly higher in mucus from patients with non-ECRS (0.11 ± 0.11 vs 0.33 ± 0.37 µg/ml, p<0.05, **Figure 3C**).

**Fig. 3.**
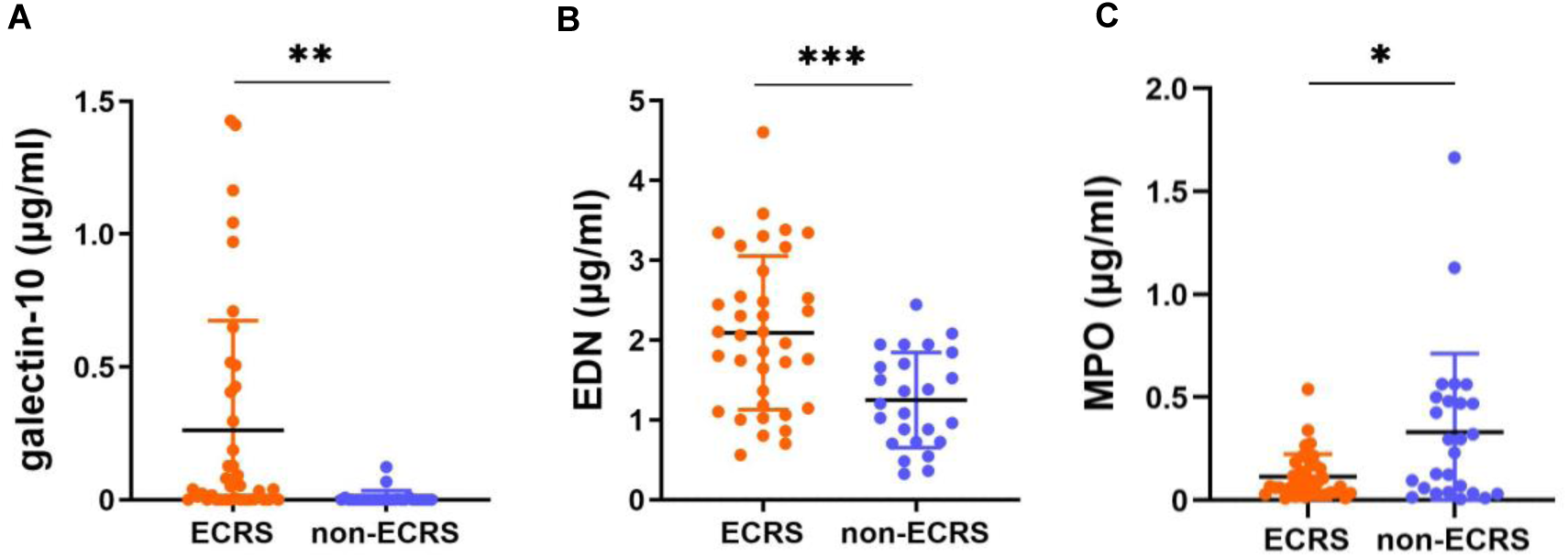
Eosinophil- and neutrophil-specific protein contents in mucus. The concentrations of **(A)** galectin-10, **(B)** eosinophil-derived neurotoxin (EDN), and **(C)** myeloperoxidase (MPO) in mucus from patients with ECRS and non-ECRS (ECRS, n=37; non-ECRS, n=25; mean ± SE, * p<0.05 ** p<0.01 *** p<0.001, Welch’s *t* test.).

The correlations between these protein levels and the physical properties or CT values of mucus were analyzed (**Table 1**). Shear viscosity was positively correlated with levels of galectin-10 and EDN, dry weight was positively correlated with levels of galectin-10, contact angle was positively correlated with levels of galectin-10 and EDN but negatively correlated with MPO, and CT values were positively correlated with levels of galectin-10 and EDN. Taken together, these results support the hypothesis that the increased eosinophils in mucus contribute to its biophysical characteristics.

**Table 1.**
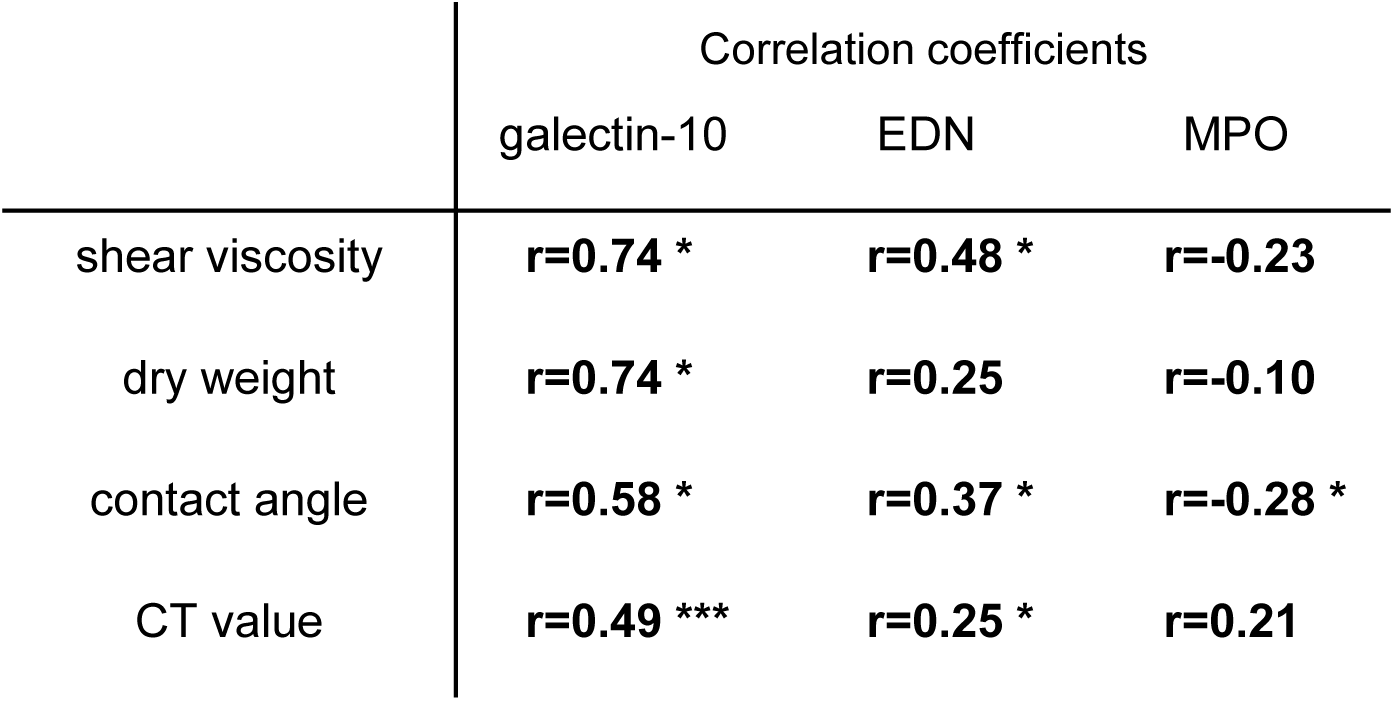
Correlations between eosinophil- or neutrophil-specific proteins in mucus and its biophysical properties. *p<0.05 ***p<0.001, Spearman’s rank correlation coefficient

|  | **Galectin-10** | **EDN** | **MPO** |
| --- | --- | --- | --- |
| **Shear viscosity** | $r = 0.74^*$ | $r = 0.48^*$ | $r = -0.25$ |
| **Dry weight** | $r = 0.74^*$ | $r = 0.25$ | $r = -0.10$ |
| **Contact angle** | $r = 0.58^*$ | $r = 0.37^*$ | $r = -0.28^*$ |
| **CT value** | $r = 0.49^{**}$ | $r = 0.25^*$ | $r = 0.21^*$ |

**Table 1. Correlation between eosinophil-specific proteins and physical properties**
|  | Correlation coefficients |  |  |
| --- | --- | --- | --- |
|  | galectin-10 | EDN | MPO |
| shear viscosity | **r=0.74 \*** | **r=0.48 \*** | **r=-0.23** |
| dry weight | **r=0.74 \*** | **r=0.25** | **r=-0.10** |
| contact angle | **r=0.58 \*** | **r=0.37 \*** | **r=-0.28 \*** |
| CT value | **r=0.49 \*\*\*** | **r=0.25 \*** | **r=0.21** |

### Aggregated ETosis cells mimic mucus properties

Unlike apoptosis, which is characterized by DNA fragmentation, activated eosinophils and neutrophils undergo ETosis to release filamentous ETs *(23, 24)*. To investigate the hypothesis that ETotic granulocytes in the mucus are involved in its biophysical properties, isolated human eosinophils and neutrophils were stimulated with phorbol 12-myristate 13-acetate (PMA) to induce ETosis and were aggregated by shear-flow (**Figure 4A**) *(25)*. The macroscopic appearance of cell aggregates showed significant differences; eosinophil aggregates (aggEETs) showed a yellow-brownish eraser-dust-like appearance, whereas neutrophil aggregates (aggNETs) showed a white gel-like appearance (**Figure 4B**). These aggregates consisted of accumulated ETs and cell debris (**Fig. S2**). The sections closely resembled clinical mucus (**Fig. 4C)**.

**Fig. 4.**
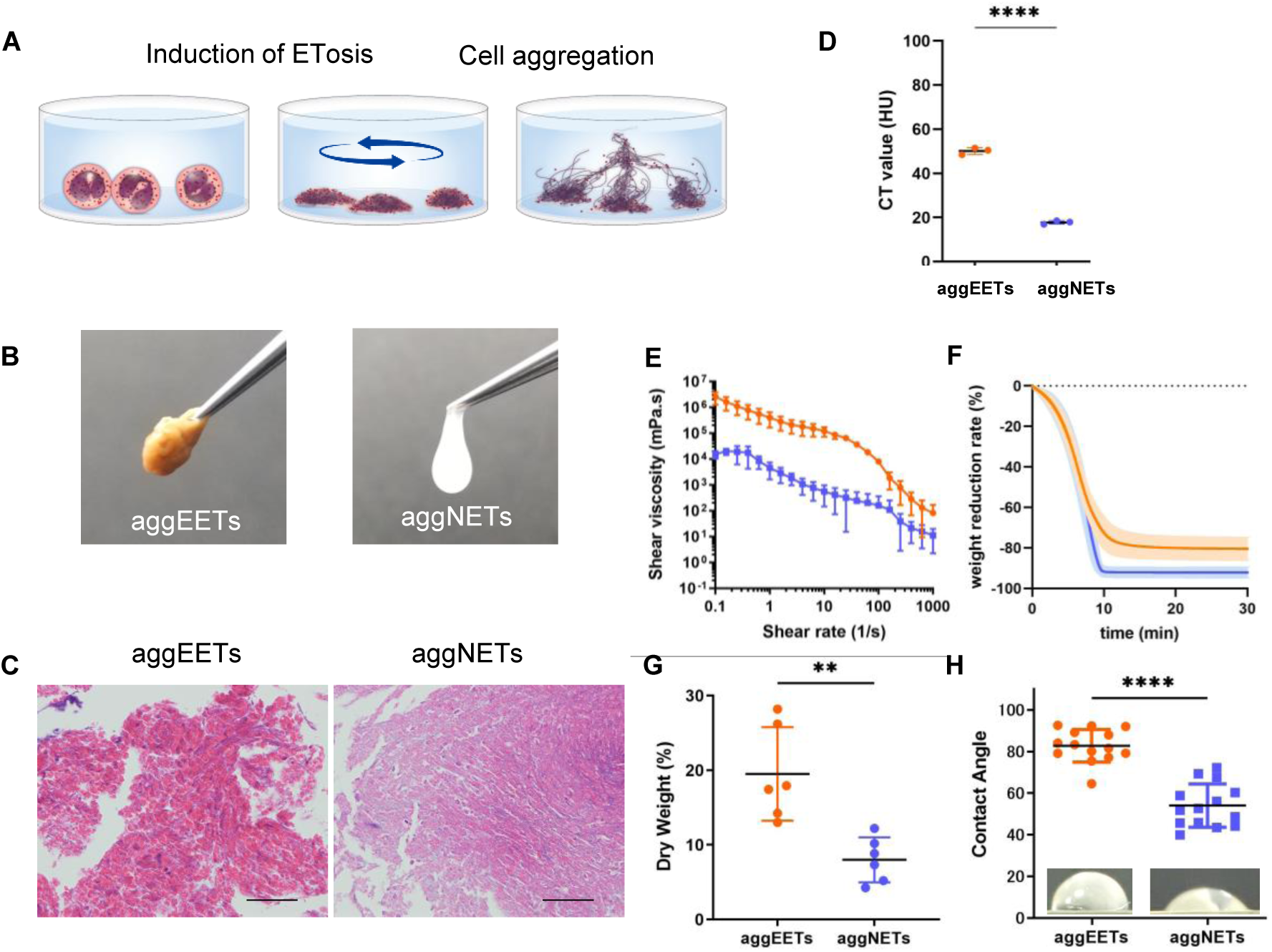
The characteristics of cell aggregates. **(A**) Preparation of aggEETs and aggNETs. Isolated human eosinophils and neutrophils were stimulated with PMA to induce ETosis. After plate shaking, aggregated cells were collected. (**B**) Macroscopic images of aggEETs and aggNETs. (**C**) Microscopic appearance of sectioned aggEETs and aggNETs (400×, hematoxylin-eosin staining, scale bar: 50 μm). (**D**) Comparison of CT values between aggEETs and aggNETs (n=3, repeated measures, Welch’s test, ****p<0.0001). (**E**) The change in shear viscosity of cell aggregates according to shear rate (orange line: aggEETs, n=3; blue line: aggNETs, n=3). (**F**) Mass reduction rate of cell aggregates using TGA (orange: aggEETs, n=6; blue: aggNETs, n=6). Lines and colored area indicate mean and ± SD, respectively. (**G**) Comparison of dry weights (aggEETs, n=6; aggNETs, n=6; *p<0.05, Welch’s test). (**H**) Comparison of contact angles (aggEETs, n=14; aggNETs, n=14; ****p<0.0001, Welch’s test).

The CT values of aggEETs were significantly higher than those of aggNETs (**Figure 4D**). Of note, the viscosity of aggEETs was higher than that of aggNETs and comparable to that of clinically obtained mucus from patients with ECRS (**Figure 4E**). In line with the mucus, the rheological properties of aggregated cells indicated a non-Newtonian shear-thinning pattern. The higher adhesion and viscosity of aggEETs were also confirmed using wall slip time-lapse images (**Movie S2**).

The weight reduction rate of aggNETs measured using the TGA was larger than that of aggEETs, and reached a plateau more quickly (**Figure 4F**). The final dry weight percentages of aggEETs and aggNETs were 19.5% ± 5.7% and 8.0% ± 2.8% (p<0.01) respectively (**Fig. 4G**). The interactions between water and polymer molecules of aggEETs and aggNETs assessed by differential scanning calorimetry *(26)* showed similar patterns (**Fig. S3**), indicating no difference in phase transition behavior and molecular mobility. The wettability assessed by the static water contact angle revealed that aggEETs showed significantly higher hydrophobicity than aggNETs (82.4° ± 2.1° vs 54.1° ± 2.8°, p<0.0001, **Fig. 4H**). These results indicate that in terms of CT value, viscosity, dry weight, and wettability, the aggEETs and aggNETs represented similar physical properties to eosinophil- and neutrophil-dominant mucus, respectively.

### Heparin enhances DNase-mediated EET degradation

EETs are composed of approximately 30-nm-diameter condensed chromatin structures and are resistant to degradation by deoxyribonuclease 1 (DNase 1), having a longer half-life than NETs *(25, 27)*. As heparin has been reported to relax NETs *(28, 29)*, we treated EETs with heparin and studied their ultrastructure. Interestingly, scanning electron microscopy revealed that heparin had a thinning effect on EETs (**Fig. 5A**). Transmission electron microscopy also revealed that the aggregated nucleosome structure of EETs was decreased after heparin treatment (**Fig. 5B**). Heparin-treated EETs consisted of smooth fibers and a globular domain, similar to the previously reported ultrastructure of NETs *(3)*. The mean diameter of non-treated EETs was 29.8 ± 0.05 µm, whereas that of heparin treated EETs was 19.2 ± 0.03 µm (**Fig. 5C**). The relaxation effect of heparin on EETs was observed within 5 minutes and was concentration dependent (**Fig. S4A**). These results indicate the disorganization of the chromatin structure, presumably through a heparin and histone interaction *(30)*. Indeed, the liquid phase interaction of histone and heparin was confirmed by the co-precipitation (**Fig. S4B, C**). Specifically, heparin-induced relaxation of EETs was canceled in the presence of excess histone (**Fig. S4D**).

**Fig. 5.**
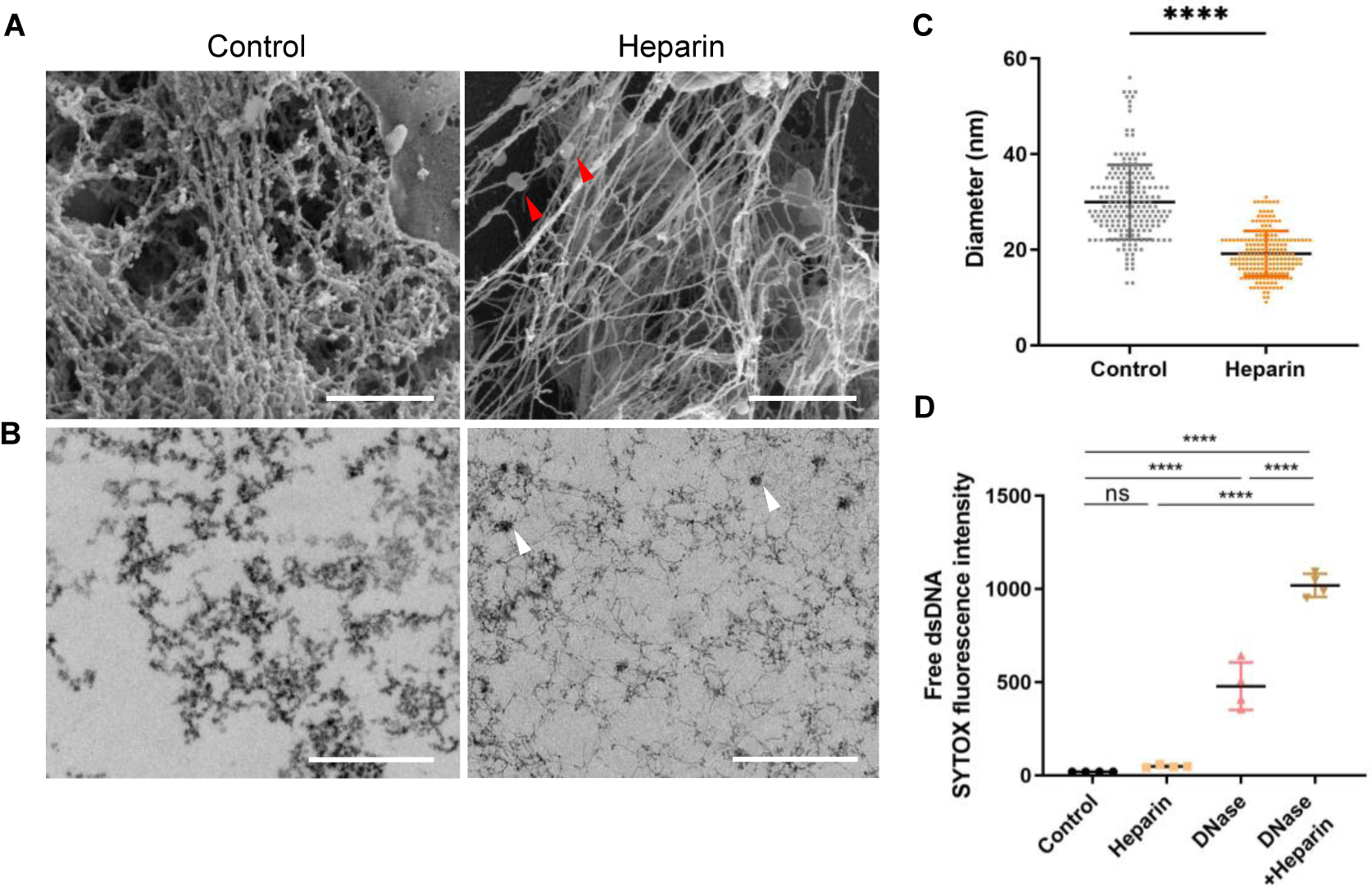
The effect of heparin and DNase on EETs. **(A)** Scanning electron microscope (SEM) images of non-treated control and heparin treated EETs (Scale bar=1 µm). The EETs consist of smooth stretch and globular domains (red arrowheads). **(B)** Transmission electron microscope images of non-treated or heparin treated EETs. White arrowheads indicate globular domain. **(C)** Comparison of fiber diameters of extracellular traps assessed by SEM (n=200, ****p<0.0001, Welch’s *t* test). **(D)** Measurement of free dsDNA detected in the culture supernatant briefly treated with heparin and/or DNase 1 (n=4, ****p<0.0001, Tukey’s multiple comparisons test).

To test whether heparin enhanced the DNase-mediated EET degradation, EETs were treated with DNase with or without heparin, and the amount of free DNA in the supernatant was quantified. Heparin itself did not increase the EET degradation, whereas DNase-induced EET degradation was significantly increased in the presence of heparin (**Fig. 5D**). These data lead us to propose that the heparin-histone interaction relaxed the chromatin structure to promote DNase-mediated DNA degradation.

### Effect of heparin and DNase on aggEETs and mucus

As aggregated cells were a complex of accumulated EETs and cell debris, we next aimed to study the effect of heparin and DNase 1 on the biophysical properties of aggEETs and aggNETs. In the static condition, we noticed that aggEETs were significantly more resistant to DNase than aggNETs, keeping their structure and DNA contents for 18 hours (**Fig. S5A**). Heparin induced a “swelling” effect on the aggEET structure and enhanced the DNase-mediated DNA degradation (**Fig. 6A, Fig. S5B**).

**Fig. 6.**
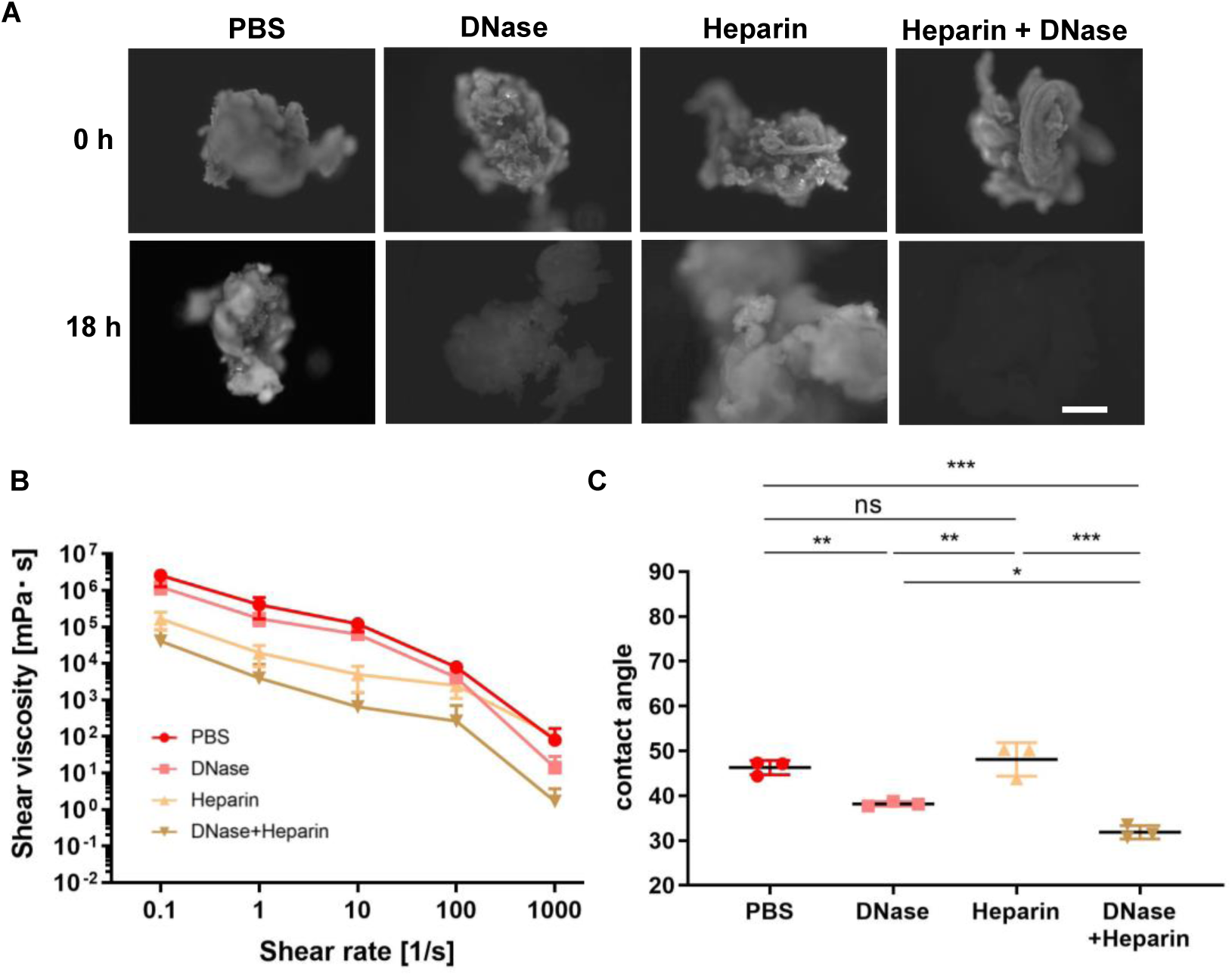
Heparin enhances the effect of DNase on aggEETs. **(A)** Morphological change in aggEETs under the effect of DNase and/or heparin was assessed by SYTOX DNA dye using inverted fluorescence microscopy (scale bar=200 µm, ×20 objective). **(B)** The cell aggregates were treated with DNase and/or heparin for 2 hours and shear viscosity was assessed by a rotational rheometer (n=3 for each group, mean ± SD). **(C)** Comparison of contact angle of cell aggregates treated with DNase and/or heparin (n=3, *p<0.05, **p<0.01, ***p<0.001, Kruskal-Wallis test: p<0.01).

The change in shear viscosity was studied by short (1 hour) exposure of aggEETs to DNase and heparin (**Fig. 6B**). Compared with the control, the most significant decrease in viscosity was observed in the aggEETs co-treated with DNase and heparin. Regarding wettability, we found that the contact angle of the aggEETs treated with DNase, but not those treated with heparin, was significantly less than that of the control. Among the groups, treatment with DNase and heparin induced the most significant decrease in hydrophobicity (**Fig. 6C**). These results indicate that aggregated DNA contributed to the hydrophobicity of aggEETs, leading to DNase-resistant viscosity.

To extend our findings to clinical samples, we examined the effect of DNase and heparin on the viscosity and wettability of mucus from patients with ECRS. As expected, eosinophilic mucin treated with DNase and heparin showed reduced shear viscosity (**Fig. 7A**). Treatment with DNase or heparin alone did not affect the wettability of eosinophilic mucin, although co-treatment with DNase and heparin significantly reduced the hydrophobicity **(Fig. 7B**). Collectively, these data indicate the therapeutic potential of heparin for facilitating DNase-induced enzymatic degradation of EETs.

**Fig. 7.**
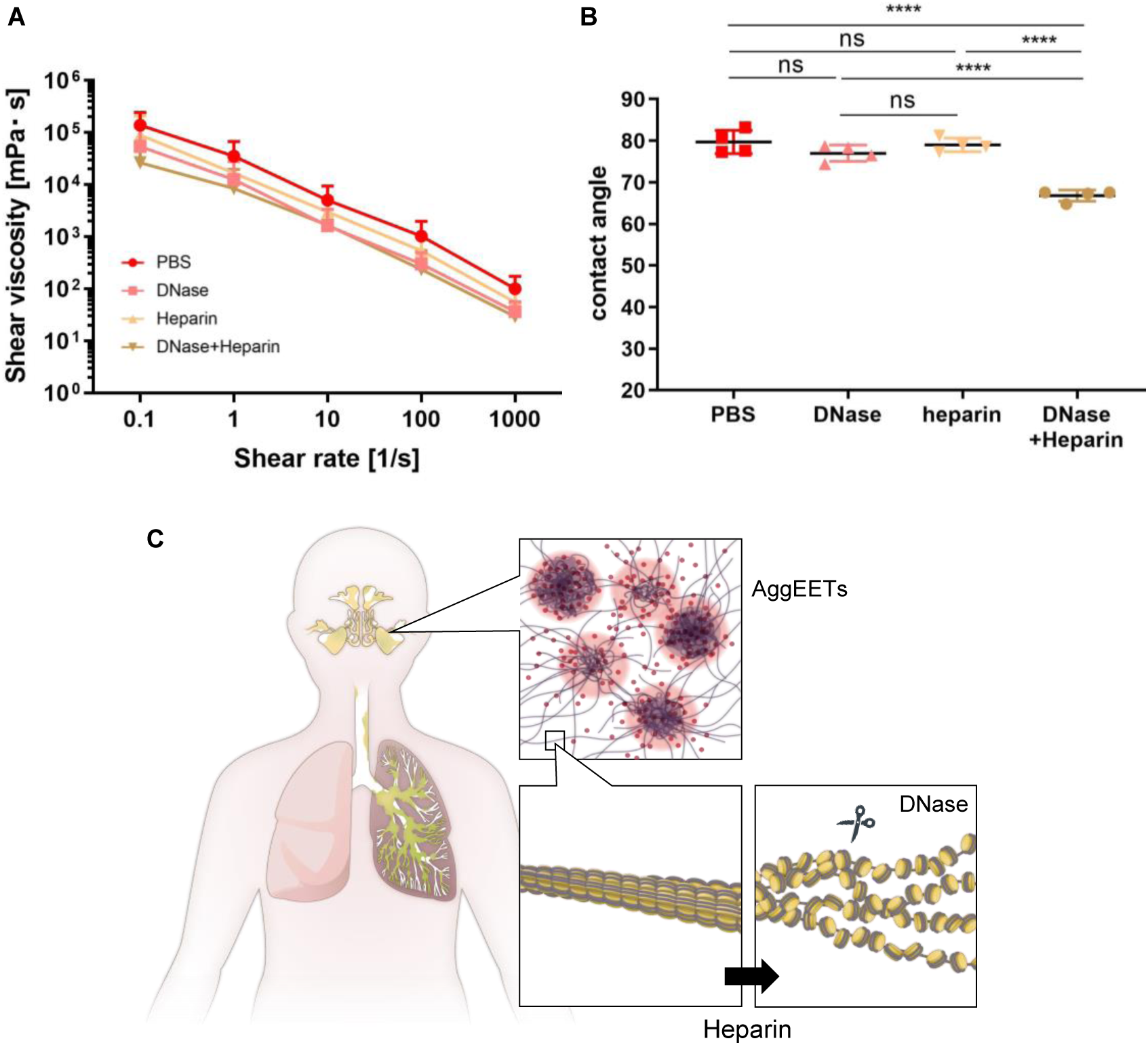
The effect of heparin on mucus in patients with ECRS. **(A)** The change in shear viscosity of mucus obtained from patients with ECRS (n=7 for each group, mean ± SD). **(B)** Comparison of contact angle of mucus treated with DNase and/or heparin (each group: n=4, ****p<0.0001, Kruskal-Wallis test: p<0.01). **(C)** Illustration of the effects of heparin on aggEETs in mucus. Airway mucus contains large amounts of aggEETs that contribute to viscosity. Heparin relaxed their condensed chromatin structure and enhanced the effect of DNase.

## DISCUSSION

The current study identified that intraluminal accumulation and activation of granulocytes contributes to the clinical properties of hyperconcentrated airway mucus and may serve as a therapeutic target. Airway mucus is a complex mixture of the following components: 1) mucus glycoprotein (mucin) derived from airway epithelial goblet cells and submucosal glands; 2) detached or migrated cell components and substances released from them; 3) serum-derived proteins such as leached albumin; 4) pathogens such as bacteria and viruses; 5) dust; 6) electrolytes; 7) substances associated with infection such as secretory IgA and other immunoglobulins, lactoferrin, lysozymes, and defensins *(31)*. Of these components, studies on the viscosity of mucus have largely focused on the hypersecretion of mucin and biological changes to it, whereas recent evidence has revealed that granulocyte-derived ET polymers contribute to the physical characteristics of inflammatory mucus *(32, 33)*.

Granulocytes are the front-line cells of innate immunity, have a short lifespan, and do not divide once they mature. They are produced in large numbers in the bone marrow and can rapidly accumulate at inflammatory sites from the blood circulation. Once entering the airways, activated granulocytes can capture pathogens by causing ETosis, and then kill them with cytotoxic protein-containing ETs *(34)*. Unlike neutrophilic inflammation, the majority of eosinophilic airway diseases are caused by sterile allergic reactions and have pathological implications. Utilizing the granulocyte-rich mucus from patients with chronic rhinosinusitis and blood-derived cells, our data indicated that the presence of aggregated ETotic eosinophils influences the biophysical characteristics of mucus. The CT values of sinus mucus were directly related to the presence of eosinophilic inflammation and its biophysical properties, supporting the utility of CT values for diagnosis and pathological evaluation in clinical practice.

Fixed airway obstruction due to eosinophilic mucus is a hallmark phenomenon of diverse eosinophilic upper and lower airway diseases. The water contact angle experiments indicated the hydrophobic properties of eosinophil-dominant mucus. Biochemically, water is the most abundant constituent in airway mucus, composing about 98% of it under normal conditions. When the water content is below 90%–93%, mucus inhibits the motility of cilia, producing mucus stasis *(35)*. In the current study, the mucus from patients with ECRS and non-ECRS was highly dehydrated (59.4% and 80.6% water, respectively), far exceeding that required to inhibit mucociliary clearance. Poor mucus clearance directly causes reduced quality of life for patients. For instance, the presence of eosinophilic mucin is reported to be associated with recurrence after endoscopic sinus surgery for chronic rhinosinusitis *(7)*. Granule proteins, cytokines, histones, and other alarmins released from inflammatory cells may cause epithelial damage and tissue remodeling when they remain intraluminally localized, generating muco-inflammatory positive feedback *(35-37)*.

To study the effects of eosinophils and neutrophils on the biophysical properties of mucus, we utilized blood-derived granulocytes. Using ETotic cell aggregates, we were able to reproduce similar physical properties as those clinically observed in eosinophil- and neutrophil-dominant mucus. While the mechanisms of inflammation have been investigated using molecular and genetic approaches, we believe that our cell population-based approach provides a better understanding of the biophysical phenomena. Although the current study focused on the properties of the mucus, a significant difference between hydrophilic aggNETs and hydrophobic aggEETs might account for formation of pathological features of diseased tissue such as neutrophilic abscess *(38)* and eosinophilic granuloma *(39)*. Our results provide a strong argument for targeting excess granulocyte accumulation and ET formation as causes of tissue dysfunction.

Aerosolized recombinant human DNase 1 (dornase alpha) is an established treatment for patients with cystic fibrosis, where it promotes the hydrolysis of NETs in airway secretions *(40, 41)*. EETs have a well-conserved chromatin structure and are more aggregated than NETs, resulting in greater stability against DNase *(25, 42)*. The current study is the first demonstration of the relaxing effect of heparin on EETs. Negatively charged heparin has a high affinity for positively charged histones, and it can disorganize chromatin fiber by liberating histones *(29, 43)*. Heparin has pleiotropic effects, such as preventing the cytotoxic effects of extracellular histone *(44)* and eosinophil granule proteins *(45, 46)*. Our data support the combined use of heparin and DNase by inhalation, endoscopic injection, or irrigation, with the potential to remove EETs to decrease the viscosity and hydrophobicity of eosinophil-dominant mucus (**Fig. 7C**). This simple mucus modification strategy could be a better approach than targeting the mucin hypersecretion pathway, which is subject to adverse consequences *(47)*.

Limitations of the current case-control study include the heterogeneity of chronic rhinosinusitis. In both ECRS and non-ECRS, chronic rhinosinusitis is classified according to the JESREC scoring system, although the pathogenesis is a mixture of inflammation by neutrophils and eosinophils in varying degrees *(48)*. Nevertheless, the concentrations of eosinophil-specific galectin-10 and EDN in mucus were associated with their properties, suggesting the importance of eosinophil density. Considering the limitations of our experimental study, we could not exclude the possibility of the presence of EET-independent granulocyte viscosity. AggEETs and aggNETs are composed of condensed cell debris including actin polymer, which may cause viscosity *(22)*. Since neutrophils contain abundant protease that may degrade organized proteins *(25)*, it is conceivable that neutrophil cell debris has a much more disintegrated structure. Other possibilities include cytolytic eosinophil production of *de novo* polymers such as Charcot-Leyden crystals *(49)*, and amyloid-like aggregation of granule protein *(4, 45, 50)*. The difference between neutrophil and eosinophil debris may be related to pathological differences, and further investigation is warranted.

In conclusion, our study revealed the characteristics of mucus and aggregated cells using radiographical, biophysical, pathological, and molecular approaches. The current study also highlights the utility of cell-population-based biophysical analysis. Intraluminal granulocyte accumulation and the activation of these granulocytes to form aggregated ETs are processes involved in the clinical features of mucus. The combination of DNase and heparin may be a promising therapeutic modality against highly viscous eosinophilic mucus.

## MATERIALS AND METHODS

### Study design

This was a case-control study comparing the CT values and physical properties of clinically obtained sinus mucus from age- and gender-matched ECRS and non-ECRS patients (**Table S1**). The sample size was based on preliminary data from our laboratory. Sinus mucus was obtained from patients with non-ECRS and those with ECRS treated by routine endoscopic sinus surgery between February 2017 and July 2019. Samples were immediately frozen at −30°C and shipped to Akita University on dry ice and stored at −80°C. Patients with ECRS were diagnosed according to the diagnostic criteria of the JESREC study score *(51)*. This scoring system assesses unilateral or bilateral disease, the presence of nasal polyps, blood eosinophilia, and dominant shadow of ethmoid sinuses on CT scans. For a definitive diagnosis of ECRS, a score of 11 or higher and a mean number of 70 or higher infiltrated eosinophil counts in sectioned tissue in the three densest areas of cell infiltration in high power fields (×400, hematoxylin-eosin staining) is required. Patients who did not meet the diagnosis of ECRS were classified as non-ECRS. Patients with an established immunodeficiency, coagulation disorder, diagnosis of classic allergic fungal sinusitis, eosinophilic granulomatosis with polyangiitis, fungal sinusitis, or cystic fibrosis, and those who were pregnant, were excluded from the study. All patients scheduled for surgery had previously failed to respond to adequate trials of conservative medical therapy administered to control their symptoms. All patients were free of systemic steroids for at least two months prior to surgery. Preoperative demographics and medical history including sex and age were obtained for each patient. For the mucus studies, the experiments were conducted with the investigator blinded to the details of the samples.

The purpose of the experimental study using mucus from patients with ECRS and cell aggregates separately dominated by neutrophils or eosinophils was to investigate the mechanism behind the highly viscous eosinophilic airway mucus with a view to developing a rationally designed treatment. For the cell-based studies, measurements were made over at least three experiments. All experiments were approved by the Akita University institutional review board, protocol approval No. 994, 1965. Written informed consent was obtained from all participants in accordance with the principles laid out in the Declaration of Helsinki.

### Cell purification

Eosinophils were isolated from peripheral blood from medication-free donors using a MACS™ system (Miltenyi Biotec, Bergisch Gladbach, Germany) with CD16-negative selection (anti-CD16 antibody-conjugated microbeads, #130-045-701, Miltenyi Biotec), as described previously *(52, 53)*. The purity of the isolated eosinophils was >98% of nucleated cells and the viability >99%. Neutrophils (>95% neutrophils, viability >98%) were obtained by positive selection using the same system. The viability and purity of the cells were analyzed via trypan blue exclusion.

### Ex vivo CT value measurement

Nasal mucus and cell aggregates were stored in microtubes and scanned three times using an Aquilion ONE CT scanner (**Fig. S1**, Cannon Medical Systems, Otawara, Japan). The scanning parameters were 120 kVp, 240 mA, 0.5-s exposure time, and 350-mm field of view. The raw data were reconstructed with an FC 21 kernel to generate 512×512 matrix images with 0.5-mm slice thickness (0.67 × 0.67 × 0.5-mm voxel resolution). Each voxel had a CT value that represented the x-ray attenuation of a substance where 0 Hounsfield units (HU) is the attenuation of water (1g/cm^3^) and −1000 HU is the attenuation of air (0 g/cm^3^). The microtubes were excluded from the obtained CT images by applying a cut-off CT threshold of −100 HU and the mucus was extracted. The CT values of the mucus were measured on each of the three scans and then averaged.

### Preparation and characterization of ETosis cell aggregates

Isolated neutrophils and eosinophils (10–100 × 10^6^ cells) were stimulated with phorbol 12-myristate 13-acetate (PMA; Sigma, 10 ng/ml) in 6-well flat-bottom tissue culture plates, in 0.3% bovine serum albumin (BSA) containing phenol red-free RPMI 1640. After 18 hours (>99% cell death with ETosis), culture plates were shaken (600 minutes^-1^, 20 minutes) with an MS1 plate shaker (IKA Works, Wilmington, NC) to induce aggregates. The aggregates were collected using a cell scraper and tweezers. In some experiments, they were fixed in 10% formalin and embedded in paraffin, followed by hematoxylin-eosin staining. For liquid phase stability, aggregates were stained with SYTOX green (1:5000) and viewed by an inverted microscope (Eclipse TE300, Nikon, Tokyo, Japan) equipped with a cooled color digital camera (Spot 1.3.0, Diagnostic Instruments, Sterling Heights, MI) in conjunction with IP Lab image analysis software (Scanalytics, Fairfax, VA). Additional characterization is provided in **Fig. S2**.

### Measurement of shear viscosity

The rheological experiments were carried out using an MCR302 rotational rheometer (Anton Paar Japan, Tokyo, Japan) with a cone–plate system (CP25-2) for shear fixation. Frequency sweep tests were performed from 0.1□rad/s to 100□rad/s. The shear thinning behavior of secretions and cell aggregates was characterized over a range of 0 to 100 s^-1^. The viscosity of the sample against shear rate was measured. All rheology studies were performed at 36°C. In some experiments, the shear viscosity was also measured with added DNase 1 (40 U/ml, New England Biolabs, Ipswich, MA) and/or heparin (300 µg/ml, Sigma-Aldrich, St. Louis, MO). DNase and/or heparin were added in sufficient quantities to soak the samples in the tubes and the tubes were incubated for one hour at 37°C. Before measurement of the shear viscosity, 20 mg of each sample was prepared.

### Thermogravimetric analysis (TGA)

A sample of each of the cell aggregates or mucus (5 mg) was placed in an alumina crucible and heated in a thermogravimetric analyzer (STA 7300, Hitachi High-Technologies, Tokyo, Japan). Samples (originally at 25°C) were heated at a rate of 10°C/minute up to a maximum of 100°C and were maintained at this temperature for 30 minutes in a stream of dry nitrogen. The percentage weight change was evaluated, and the final weight was considered as the dry weight.

### Water contact angle analysis

Mucus or cell aggregates were uniformly smeared onto a slide and dried in ambient air for 6 hours. For measurement of the contact angle, a 4-µl water droplet was placed using a manipulator and imaged using a digital camera 30 s after the release of the droplet. The contact angle values of the droplets were recorded using the ImageJ contact angle plug-in (https://imagej.nih.gov/ij/). In some experiments, we also measured the contact angle of samples treated overnight with DNase 1 (40 U/ml) and/or heparin (300 µg/ml, Sigma-Aldrich, St. Louis, MO). Prepared samples were smeared onto a slide and the contact angle was measured as described above.

### Measurement of the levels of EDN, galectin-10, and MPO

Ten microliters of 0.1% dithiothreitol were added to 10-mg mucus samples, which were then stirred for 15 minutes before being quickly frozen in liquid nitrogen. The samples were crushed with a multi-bead shocker (Yasui Kikai, Osaka, Japan) at 250 bpm for 10 s and then with an ultrasonic crusher (Bioruptor, BM Equipment Co., Ltd. Tokyo, Japan). The supernatant was collected by centrifugation (10 000× g, 10 minutes). The levels of EDN, galectin-10, and MPO were measured using ELISA assay kits (EDN: 7630, MBL, Nagoya, Japan; galectin-10: Cloud-Clone Corp., Katy, TX, MBL, Aichi, Japan; MPO: Enzo Life Sciences Inc, Farmingdale, NY).

### Scanning electron microscopy (SEM)

Eosinophils were added to round coverslips in culture plates and stimulated with PMA (10 ng/ml) for 3 hours or more to induce EET formation, with or without heparin (300 µg/ml, 30 minutes). They were then immediately fixed in a mixture of freshly prepared aldehydes (1% glutaraldehyde and 1% paraformaldehyde) in 0.1 M phosphate buffer (pH 7.4) for 1 hour at room temperature and processed for SEM. Coverslip-adherent cells were postfixed with 1% osmium tetroxide in water for 1 hour and dehydrated in an ascending ethanol series from 50% (vol/vol) to absolute ethanol (10 minutes per step). Cells were then critical point dried in carbon dioxide. Coverslips were mounted on aluminum holders, sputtered with 5-nm gold, and analyzed in a scanning electron microscope (Thermo Fisher Scientific FEI Quanta 3D FEG Dual Beam (SEM/FIB)], which enabled excellent high resolution surface imaging.

Qualitative analyses were performed on 70 electron micrographs (n=35 for each group) at different magnifications. For quantitative analyses of EETs, a total of 20 electron micrographs from cytolytic eosinophils showing EETs (10 from untreated and 10 from heparin-treated cells) were analyzed at 80 000×. The diameters of 20 EET fibers randomly selected from each electron micrograph were measured, with a total of 400 fibers being analyzed (n=200 fibers in each group). The mean diameter of the EET fibers was then determined for each group.

### Transmission electron microscopy (TEM)

Eosinophils were seeded on Aclar film (Nishin EM, Tokyo, Japan), stimulated with 10 ng/ml of PMA for 3 hours, and then immediately fixed in a mixture of freshly prepared aldehydes (1.25% glutaraldehyde and 1% paraformaldehyde) in 0.1 M cacodylate buffer. After incubation for 2 hours in 0.1 M cacodylate buffer containing 1% OsO_4_, the specimens were dehydrated in an ethanol series, passed through propylene oxide, and embedded in epoxy resin. Ultrathin sections (80 nm) were collected on copper grids and stained for 20 minutes in 4% uranyl acetate and 0.5% lead citrate. The specimens were viewed with a Hitachi H-7650 transmission electron microscope at 100 kV.

### EET and aggEET dissolution assays

Isolated eosinophils were induced to ETosis by stimulation with PMA (10 ng/ml) for 18 hours. The medium was replaced with 100 µl of Ca^2+^-containing Hank’s balanced salt solution with Picogreen (Thermo Fisher Scientific, Waltham, MA, 1:100) and incubated with or without DNase 1 (20 U/ml) and heparin (5 mg/ml). The plate was shaken gently (400 rpm) for 5 minutes at room temperature. After pipetting, 100 µl of medium was aspirated and centrifuged (500× g, 5 minutes). The fluorescence intensity of the supernatant was immediately measured to quantify the free DNA using a GloMax-Multi detection system (Promega, Madison, WI). In the static condition, aggEETs were incubated at 37°C (0.3% BSA/RPMI medium) in the presence of DNase 1 (40 U/ml). To visualize DNA, SYTOX was added to the medium. Fluorescent images were obtained at indicated time points under an inverted microscope (Eclipse TE300, Nikon, Tokyo, Japan) equipped with a cooled color digital camera.

### Statistical Analysis

Results are expressed as mean ± SD. The data analysis in this study was performed using GraphPad Prism software, Version 7 (GraphPad, San Diego, USA). The Wilcoxon signed rank test or Welch’s *t* test were used to evaluate differences between two groups. Multiple comparison tests were used to compare means between groups. Correlation analysis was performed using Spearman’s rank correlation. A *p*-value of <0.05 was considered statistically significant.

## Data Availability

All data produced in the present study are available upon reasonable request to the authors

## List of Supplementary Materials

**Fig. S1**. Computed tomography (CT) imaging of mucus and cell aggregate samples

**Fig. S2**. Structures of aggregated eosinophil extracellular traps (aggEETs) and aggregated neutrophil extracellular traps (aggNETs)

**Fig. S3**. Differential scanning calorimetry (DSC) chart of aggEETs and aggNETs

**Fig. S4**. Heparin-induced relaxation ability on EETs

**Fig. S5**. Stability of aggEETs and aggNETs against DNase

**Table S1**. Clinical characteristics of patients with eosinophilic chronic rhinosinusitis (ECRS) and non-eosinophilic chronic rhinosinusitis (non-ECRS).

**Movie S1**. Surgical removal of sinus mucus

**Movie S2**. Wall slip time-lapse images

## Acknowledgments

The authors are grateful to Noriko Tan, Satomi Misawa, Nozomi Tanaka, and Chikako Furutani for their outstanding technical assistance. We thank Satoshi Marumo for helpful discussion and Edanz (https://jp.edanz.com/ac) for editing a draft of this manuscript.

## Funding

AstraZeneca Evidence Connect Externally Sponsored Research (SU)

Charitable Trust Laboratory Medicine Research Foundation of Japan (SU)

JSPS KAKENHI 15KK0329, 20K08794, 21K08434, and 21K07833 (SU)

JSPS KAKENHI 17K09993 (MT)

JSPS KAKENHI 17K17611 (YK).

JSPS KAKENHI 22K08598 (IM)

JSPS KAKENHI 17K11356, 25293348, and 20H03832 (TY)

Research Grant on Allergic Disease and Immunology from the Japan Agency for Medical

Research and Development JP22ek0410097 (KA)

Conselho Nacional de Desenvolvimento Científico e Tecnológico 406019 and 309734 (RCNM)

Fundação de Amparo à Pesquisa do Estado de Minas Gerais (FAPEMIG) (RCNM)

## Author contributions

Conceptualization: YM, SU

Methodology: SU, YM, MF, MJ, NT, RCNM

Investigation: YM, MF, HT, YN, MJ, YM, MA, YK, YM, NT, HS, YS, NO, JT, AM, TY, RCNM, SU

Visualization: YM, YM

Funding acquisition: SU, MT, YK, TY, KA, IM, RCNM

Project administration: SU

Supervision: TH, KA, PFW, SU

Writing – original draft: YM, YN, MF, SU

Writing – review & editing: IM, MJ, TY, RCNM, PFW

## Data and materials availability

All data are available in the main text or the supplementary materials.

