## Supplementary materials for "Aggregated eosinophils characterize airway mucus properties"

**Supplementary Figures**

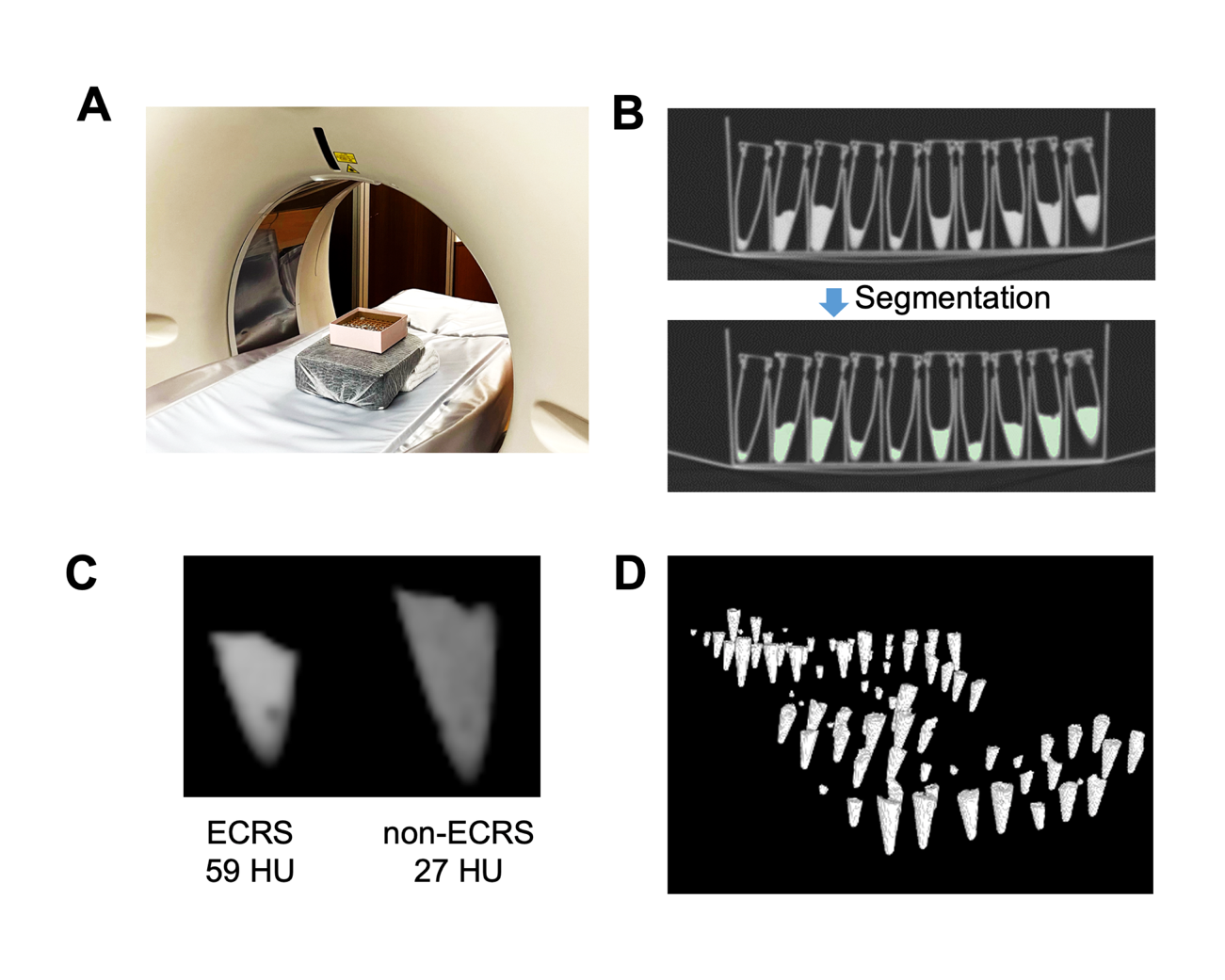

**Fig. S1. Computed tomography (CT) imaging of mucus and cell aggregate samples**

**(A)** Surgically obtained mucus and cell aggregate samples were assessed using ultra-high resolution CT scanning. **(B)** The sample in each microtube was segmented by thresholding with a cut-off of CT value of −100 Hounsfield units (HU). **(C)** Representative images of segmented mucus from patients with ECRS and non-ECRS. Mean CT values are indicated. **(D)** Three-dimensional representations of samples on CT.

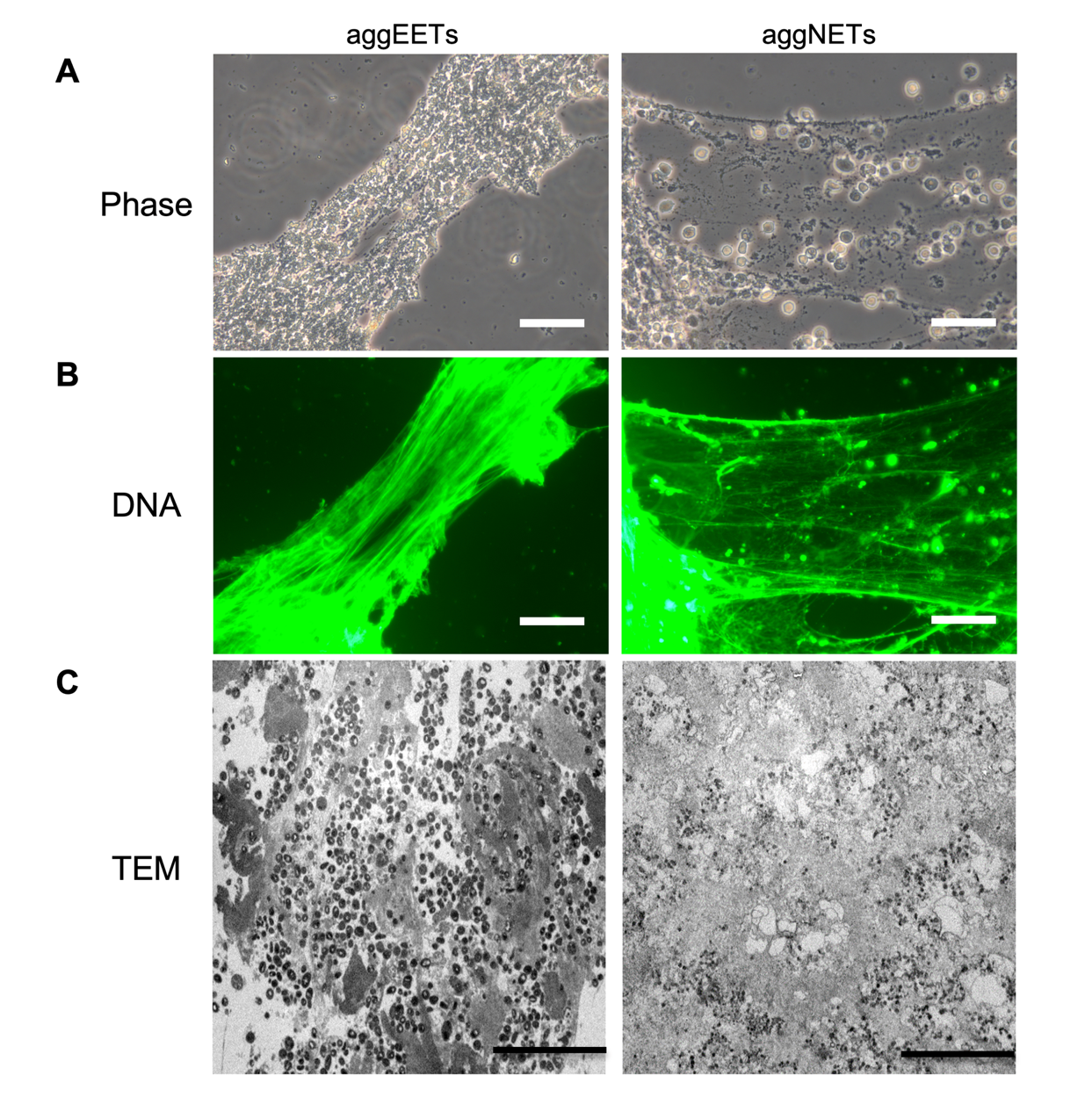

**Fig. S2. Structures of aggregated eosinophil extracellular traps (aggEETs) and aggregated neutrophil extracellular traps (aggNETs).**

**(A)** To induce ETosis (extracellular trap cell death), isolated eosinophils and neutrophils were stimulated with phorbol 12-myristate 13-acetate (PMA; 10 ng/ml) overnight in 0.3% BSA containing phenol red-free RPMI 1640 and cell aggregates were prepared as described in the Materials and Methods. Cell aggregates were stained with the cell-impermeable fluorescent DNA dye SYTOX green and smeared onto a glass slide. Phase contrast microscopic images were captured using an inverted microscope (DMI 4000B, Leica, Tokyo, Japan). **(B)** Fluorescence images of SYTOX green-stained aggregated extracellular traps. DNA fibers of aggEETs are denser than those of aggNETs. Scale bar = 50 µm. **(C)** Transmission electron microscopic (TEM) images show cell debris including extracellular granules and disintegrated nuclear contents. Scale bar = 10 µm.

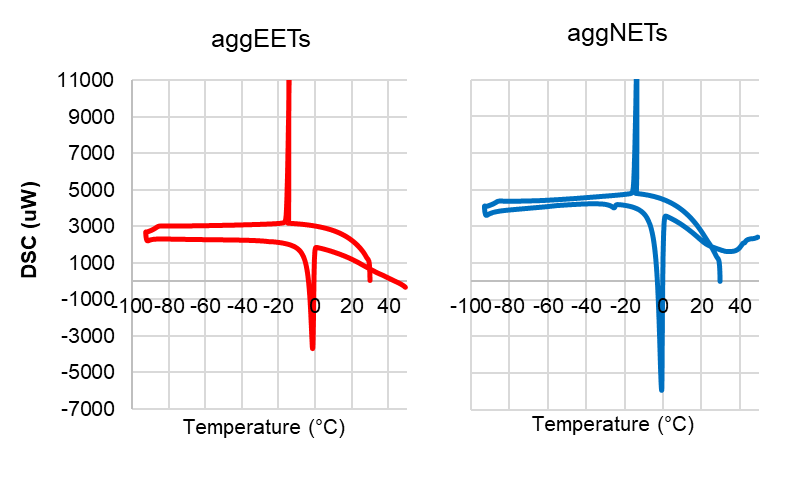

**Fig. S3. Differential scanning calorimetry (DSC) chart of aggEETs and aggNETs.**

To evaluate the structure of water in cell aggregates, the thermal behavior was measured using a DSC-7000X (Hitachi High-Technologies) calorimeter. Shown is a DSC chart of neutrophil and eosinophil derived aggregates that were cooled from 30°C to −100°C and then heated from −100°C to 50°C (2.5°C/minute). Five mg of the aggNETs and aggEETs were placed in aluminum pans. The thermograms were acquired between 30°C and −100°C during cooling, and between -100°C and 50°C during heating, at a rate of 2.5°C/minute in a stream of nitrogen (30 ml/minute). The melting peak can be seen around 0°C, indicating that the water contained in the sample was free water. The absence of a cold crystallization peak around −40°C in the heating curve can be noted, indicating the absence of intermediate water.

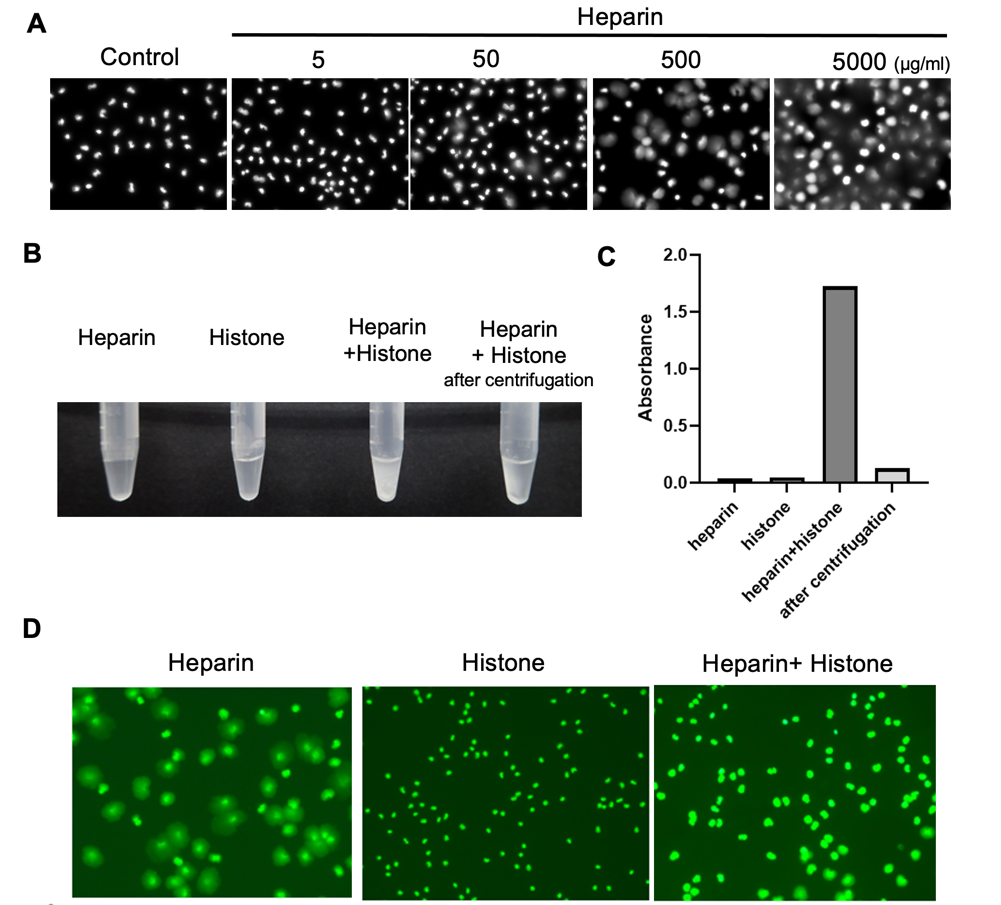

**Fig. S4. Heparin-induced relaxation ability on EETs**

**(A)** Isolated eosinophils (1 x 10^5^) were stimulated with PMA (10 ng/ml) in 0.3% BSA/RPMI 1640 medium for 3 hours to induce ETosis. The spread of extracellular DNA was visualized using the cell-impermeable DNA dye SYTOX (1:5000). Heparin was then added to the medium in the indicated concentrations and the medium was incubated for 5 minutes. The fluorescent images were obtained using an inverted microscope (Eclipse TE300, Nikon, Tokyo, Japan) equipped with a cooled color digital camera. EET decondensation was observed in a concentration-dependent manner. **(B)** Heparin (50 µg/ml) and/or histone (2.5 mg/ml) in PBS showed a clear solution, but the mixture (incubated at 37°C for 30 minutes) showed a turbid appearance. Centrifugation (14 000 x g, 5 minutes) lead to the precipitation of the heparin-histone complex. **(C)** The absorbance of the supernatants of the indicated solution was measured at 380 nm using a FLUOstar Omega system (BMG Labtech, Offenburg, Germany). **(D)** Heparin-induced relaxation of EETs was canceled by the presence of the excess histone. Isolated eosinophils were incubated with PMA for 3 hours and histone (2.5 µg/ml) was added to the medium and the medium incubated for 5 minutes at 37°C. Heparin (50 µg/ml) was reacted for 30 minutes and fluorescent images were captured using an inverted microscope (DMI 4000B; Leica, Tokyo, Japan) equipped with a cooled color digital camera (DFC450C; Leica).

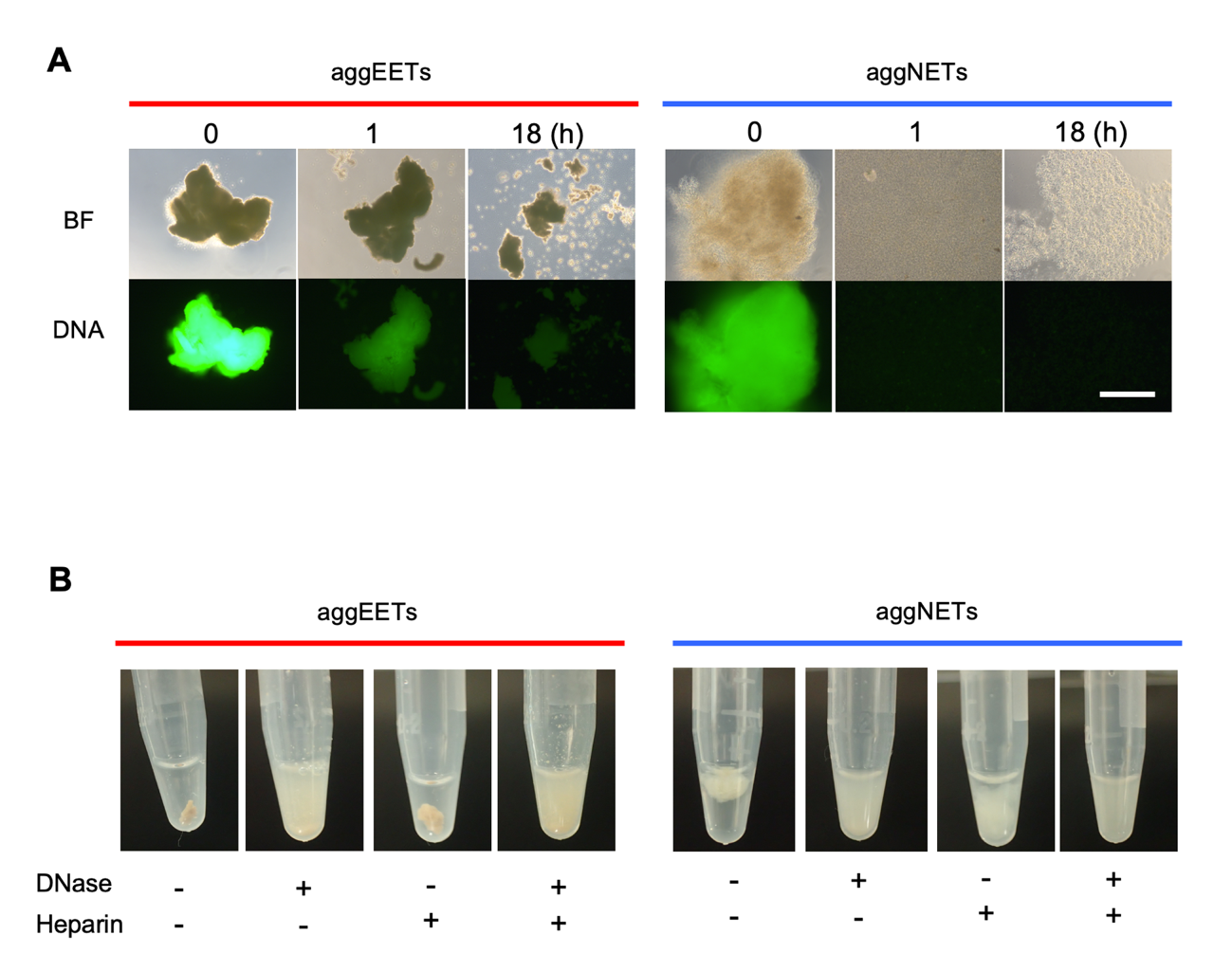

**Fig. S5. Stability of aggEETs and aggNETs against DNase**

**(A)** In the static condition, aggEETs and aggNETs were incubated at 37°C (0.3% BSA/RPMI medium) in the presence of DNase 1 (40 U/ml). To visualize DNA, SYTOX was added to the medium. Bright field (upper panels) and fluorescent images (lower panels) were obtained at the indicated time points using an inverted microscope (DMI 4000B, Leica, Tokyo, Japan). In contrast to rapid degradation of aggNETs, the structure and SYTOX-signal of aggEETs remained stable over time, indicating resistance against DNase. Scale bar = 400 µm. **(B)** Macroscopic appearance of aggEETs and aggNETs treated with DNase 1 (40 U/ml) and/or heparin (300 µg/ml) for 2 hours at 37°C followed by shaking for 10 seconds.

**Supplementary table**

| **Clinical characteristics** | **ECRS (n=40)** | **non-ECRS (n=27)** | **P Value** |
| --- | --- | --- | --- |
| **Age (y)** Mean ± SD | 44.8 ± 7.9 | 45.9 ± 12.5 | NS |
| **Sex Male,** No. (%) | 13 (33) | 13 (48) | NS |
| **Female,** No. (%) | 27 (67) | 14 (52) |  |
| **Asthma,** No./ total (%) | 35/40 (87) | 6/27 (22) | p<0.0001 |
| **Drug allergy,** No./ total (%) | 19/40 (48) | 0/24 (0) | p<0.0001 |
| **Blood eosinophil (%)** Mean ± SD | 11.0 ± 4.9 | 5.1 ± 3.4 | p<0.0001 |
| **Tissue eosinophil count** Median (range) | 100 (70–333) | 1.3 (0–38) | p<0.0001 |

**Table S1. Clinical characteristics of patients with eosinophilic chronic rhinosinusitis (ECRS) and non-eosinophilic chronic rhinosinusitis (non-ECRS).**

**Supplementary movies**

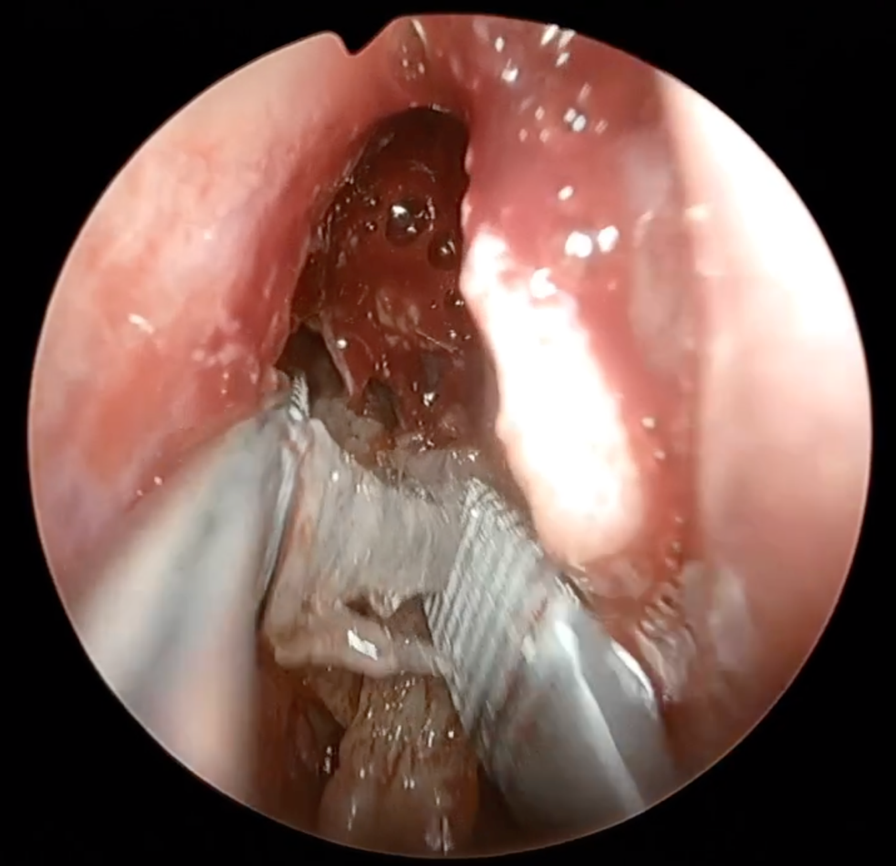

**Movie S1. Surgical removal of sinus mucus**

Eosinophilic mucin was removed from the sinus of a patient with ECRS during endoscopic sinus surgery.

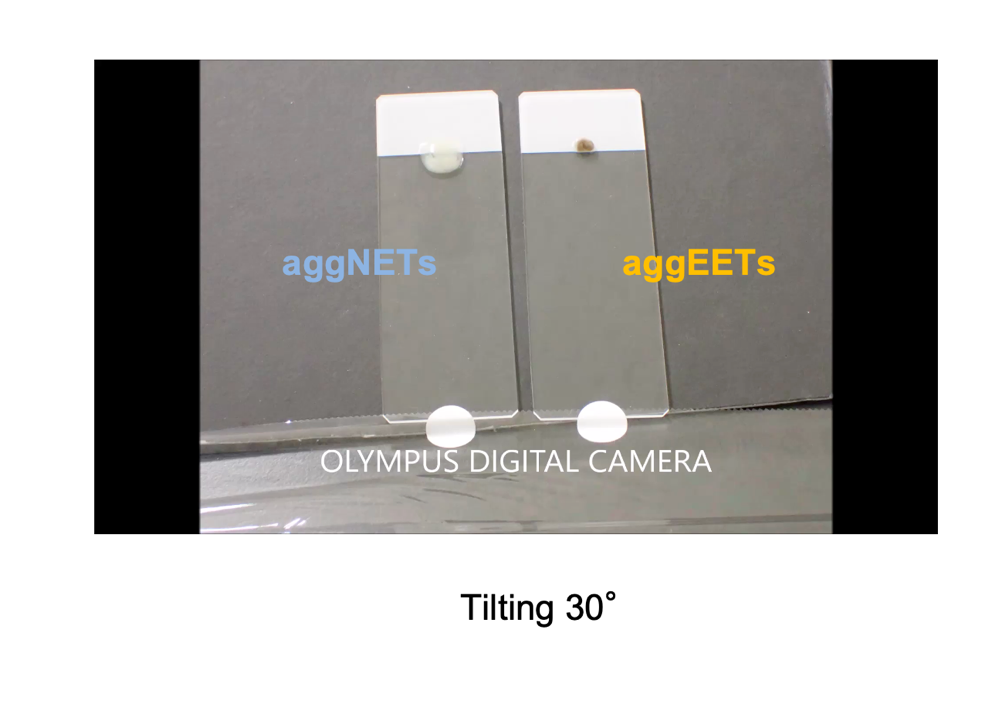

**Movie S2. Wall slip time-lapse images**

To confirm differences in viscous resistance between aggEETs and aggNETs (4 × 10^7^ cells), a wall slip test that reflects the fluid’s viscous resistance was conducted. The samples were placed on a glass slide inclined at 30 degrees. Spontaneous sliding was photographed every 30 seconds and images were captured with a digital camera (TG4BLK; ORYMPUS, Tokyo, Japan).
